# Association of ABO blood group with COVID-19 severity, acute phase reactants and mortality

**DOI:** 10.1101/2021.04.19.21255738

**Authors:** Uzma Ishaq, Asmara Malik, Jahanzeb Malik, Asad Mehmood, Azhar Qureshi, Talha Laique, Syed Muhammad Jawad Zaidi, Muhammad Javaid, Abdul Sattar Rana

## Abstract

**Background and objective:** The ABO blood group system has been associated with infectious and noninfectious disease, including dengue, hepatitis B virus (HBV), and severe respiratory syndrome coronavirus (SARS), etc. Coronavirus disease 2019 (COVID-19) is the ongoing pandemic with multitude of manifestations and association of ABO blood group in South-East Asian population needs to be explored.

**Methods:** It was a retrospective study of patients with real time polymerase chain reaction (RT-PCR) diagnosis of COVID-19 at Advanced Diagnostics and Liver Center between April 2020 to January 2021. Blood group A, B, O, and AB were identified in every participant, irrespective of their RH type and allotted groups 1, 2,3, and 4, respectively. Cox regression and logistic regression were used for inferential statistics.

**Results:** The cohort included 1067 patients: 521 (48.8%) of blood group O, 295 (27.6%) of blood group B, 202 (18.9%) of blood group A, and 49 (4.5%) of blood group AB. The majority of the patients were males 712 (66.7%) with an average body mass index (BMI) of 27.45 ± 3.53. Patients with AB blood group stayed a median (IQR) of 14 (5, 27) days while A blood group cohort stayed 13 (6,27) days and overall 10.6% COVID-19-related mortality was observed at our center, with 13.9% in blood group A as the majority of COVID-19 deaths. Regarding severity of COVID-19 disease, there was a trend towards critical disease in blood group A and O (n=83, 41.1%; n=183, 35.1%; OR, 11.34 (95% CI, 46.79-53.22); p<0.001). Logistic regression demonstrates blood group O and AB as predictors for severe COVID-19 disease (O: OR: 0.438 (95% CI: 0.168-1.139) p=0.090; AB: OR: 0.415 (95% CI: 0.165-1.046) p=0.062) and cause-specific hazards ratio (HR) for survival function was 3.206 (p=0.361) among all blood groups.

**Conclusion:** Although the prevalence of blood group O was higher in this cohort, hospital stay, severity of disease, and mortality were associated with blood group A. Further studies are needed for understanding the underlying mechanism behind the association of blood groups with COVID-19.

## Introduction

Coronavirus disease 2019 (COVID-19) represents a public health emergency causing economic and health care system collapse worldwide. As of April 2021, approximately three million people have been diagnosed with COVID-19 with the numbers increasing exponentially [1].

With some of the regions surviving through the third wave of the pandemic, there has been a multitude of manifestations associated with COVID-19, including the cardiovascular, respiratory, and gastrointestinal systems [2,3,4].

Apart from documentary evidence of auto-antigenicity and reporting of hematological complications, many studies have investigated the pathway to viral entry into the human hosts [5,6]. One such hypothesis resides in the ABO blood group and its association with the severity of disease in Hepatitis B Virus (HBV), Middle-Eastern Respiratory Syndrome Coronavirus (MERS), and Severe Acute Respiratory Syndrome Coronavirus (SARS) [7]. Several investigations have reported an association between the ABO blood group and COVID-19 [8]. A study on COVID-19 demonstrated a cross-replicating association signal at locus 9q34.2, which coincides with the ABO blood group locus [9]. However, the association between the blood groups and the severity of the disease is still unclear. Several studies have shown an increased risk of infectivity with blood group A while conferring a low risk of COVID-19 infection with blood group O [10]. In South-East Asia however, blood group O and B is prevalent and studies demonstrating the severity of COVID-19 disease and COVID-19-associated mortality in this population subset were lacking [11]. Hence, we conducted a retrospective analysis on a cohort of COVID-19 patients, analyzing their ABO blood types, and observed overall COVID-19-associated mortality and severity of disease in association with the blood type.

## Methods

This was a retrospective investigation conducted at Advanced Diagnostics and Liver Center after approval from the ethical review board of our institute (ID: ADC/17/20) according to the Declaration of Helsinki. All participants or their guardians gave written informed consent before data collection. A total of 1067 COVID-19 patients were included in this study from April 2020 through January 2021 and the demographic data, comorbid conditions, epidemiological data, laboratory tests, hospital stay, and mortality rates were extracted via the electronic system of our institute. Every patient was confirmed positive SARS-COV-2 via real-time reverse transcription-polymerase chain reaction (RT-PCR). The severity of COVID-19 was classified into, mild, moderate, and critical according to the Centers for Disease Control (CDC) guidelines. Mild cases were classified as normal oxygen saturation on room air and mild severity of symptoms of COVID-19. Oxygen saturation of < 90% and respiratory rate of > 30 breaths/min or < 50% lung involvement on high resolution computed tomography (HRCT) were labeled as moderate and those requiring mechanical ventilation were labeled as critical. Patients with known hemoglobinopathies or other blood disorders were excluded.

All the tests were carried out in the clinical laboratory of Advanced Diagnostics under the standard procedures according to Punjab Health Commission. Hemoglobin (Hgb), and white blood cells (WBC) were performed on an XN-3100 Sysmex hematology analyzer. C-reactive proteins and D-dimers were analyzed on Cobas® c3011 analyzer (Roche Diagnostics) and interleukin-6 (IL-6), and Procalcitonin was analyzed via electrochemiluminescent immunoassay (ECLIA) in the Elecsys® 2010 immunoassay system. ABO blood group and the cross-match were done manually by an expert hematologist (UI). Group A, B, O, and AB were identified in every participant, irrespective of their RH type and allotted groups 1, 2,3, and 4 respectively. Statistical analysis was carried out with Statistical Package for the Social Sciences (SPSS) version 26 (IBM Corp, Armonk, NY, USA). After normality adjustments using the Wilk-Shapiro test, quantitative variables were presented as mean ± standard deviation (SD) for normal distribution and median (Interquartile range: IQR) for non-normal distribution. Qualitative variables were presented as frequency and percentages. Comparison of the four groups was analyzed by Student’s t-test and Chi-square test was used for qualitative variables. For outcomes assessment, cumulative incidence and Kaplan Meier curves were plotted for all four groups against the number of days in the hospital and mortality rate. Odds ratio (OR) and 95% confidence interval (CI) were analyzed for the severity of COVID-19 disease, mortality, and hospital stay. Correlation between blood group and lab parameters was presented as histogram distributed among the four groups. Logistic regression was done for predictors of severe COVID-19 disease for ABO blood groups. A p-value of less than 0.05 was considered significant.

## Results

A total of 1067 patients were admitted to Advanced Diagnostics and Liver Center from April 2020 through January 2021 who complied with our study criteria for analysis. The mean age of the patients in group 1, 2, 3, and 4 was 47.37 ± 20.21, 47.71 ± 18.45, 47.54 ± 18.84, and 48.87 ± 21.31 years, respectively. The majority of the patients were males 712 (66.7%) with an average body mass index (BMI) of 27.45 ± 3.53. Overall, one-quarter of the patients were diabetic 310 (29%), had hypertension 361 (33.8%), cardiovascular disease (CVD) 282 (26.4%), and 96 (8.9%) had chronic kidney disease (CKD). The majority of blood groups in descending order for this cohort were O (48.8%), B (27.6%), A (18.9%), and 4.5% were AB blood type.

The average median (IQR) hospital stay was 12 (6,25) days. Patients with AB blood group stayed a median (IQR) of 14 (5, 27) days while A blood group cohort stayed 13 (6,27) days. It was statistically non-significant between all four groups. There was overall 10.6% COVID-19-related mortality at our center, with 13.9% in blood group A as the majority of COVID-19 deaths. However, this did not reach statistical significance. Regarding severity of COVID-19 disease, there was a trend towards critical disease in blood group A and O (n=83, 41.1%; n=183, 35.1%; OR, 11.34 (95% CI, 46.79-53.22); p<0.001). Baseline characteristics, patient demographics, the severity of COVID-19, total hospital stay, and mortality rates are presented in **Table 1**. The overall incidence of the ABO blood type in the COVID-19 cohort and cumulative hazard of cause-specific death is demonstrated in **Figure 1** and the frequency of acute phase reactants (IL-6, CRP, Procalcitonin, D-dimers) with ABO blood type is exhibited in **Figure 2**.

**Table 1.** Patient demographics and baseline characteristics of COVID-19 patients with different blood groups. Normally distributed variables expressed as mean ± SD, abnormally distributed variables expressed as median (IQR). Categorical variables presented as n (%). P < 0.05 as significant. Standard deviation (SD), interquartile range (IQR), body mass index (BMI), diabetes mellitus (DM), hypertension (HTN), cardiovascular disease (CVD), chronic kidney disease (CKD), hemoglobin (Hgb), white blood count (WBC), C-reactive protein (CRP), interquartile range (IQR), standard deviation (SD), odds ratio (OR), confidence interval (CI)

| **Variable** | **Blood Group**<br>**A (n=202)** | **Blood Group**<br>**B (n=295)** | **Blood Group**<br>**O (n=521)** | **Blood Group**<br>**AB (n=49)** | **OR**<br>**(95%**<br>**CI)** | **P-value** |
| --- | --- | --- | --- | --- | --- | --- |
| **Age, years,**<br>**mean ± SD** | 47.37 ± 20.21 | 47.71 ± 18.45 | 47.54 ± 18.84 | 48.87 ± 21.31 | - | 0.444 |
| **Gender, n(%)** |  |  |  |  |  | 0.342 |
| Male | 138 (62.4%) | 239 (69.3%) | 283 (67.5%) | 52 (63.4%) | - |  |
| Female | 83 (37.6%) | 106 (30.7%) | 136 (32.5%) | 30 (36.6%) | - |  |
| **BMI, kg/m<sup>2</sup>,**<br>**mean ± SD** | 27.34 ± 3.48 | 27.72 ± 3.67 | 27.35 ± 3.76 | 27.48 ± 3.22 | - | 0.966 |
| **Comorbid**<br>**conditions,**<br>**n(%)** |  |  |  |  |  |  |
| DM | 67 (30.3%) | 97 (28.1%) | 119 (28.4%) | 27 (32.9%) | - | 0.744 |
| HTN | 80 (36.2%) | 106 (30.7%) | 146 (34.8%) | 29 (35.4%) | - | 0.391 |
| CVD | 61 (27.6%) | 102 (29.6%) | 95 (22.7%) | 24 (29.3%) | - | 0.407 |
| CKD | 20 (9%) | 33 (9.6%) | 38 (9.1%) | 5 (6.1%) | - | 0.985 |
| **COVID-19**<br>**severity, n(%)** |  |  |  |  |  |  |
| Mild | 51 (25.2%) | 82 (27.8%) | 156 (29.9%) | 30 (61.2%) | 6.27<br>(9.09-9.77) | **<0.001** |
| Moderate | 68 (33.7%) | 128 (43.4%) | 182 (34.9%) | 14 (28.6%) | 4.88<br>(19.98-21.55) | **0.036** |
| Critical | 83 (41.1%) | 86 (29.2%) | 183 (35.1%) | 5 (10.2%) | 11.34<br>(46.79-53.22) | **<0.001** |
| **Hospital stay,**<br>**median(IQR)** | 13 (6,27) | 12 (6,25) | 12 (5,24) | 14 (5,27) | 3.78<br>(44.03- | 0.364 |
|  |  |  |  |  | 50.18) |  |
| **Mortality,**<br>**n(%)** | 28 (13.9%) | 28 (9.5%) | 52 (10%) | 5 (10.2%) | 2.38<br>(30.28-34.71) | 0.412 |
| **Lab**<br>**parameters,**<br>**median(IQR)** |  |  |  |  |  |  |
| Hgb (g/dl) | 12.7<br>(10.9,13.8) | 12.3<br>(10.7,14.1) | 12.7<br>(11.2,14.8) | 12.7 (11.1,15) | - | 0.377 |
| WBC (mm<sup>3</sup>) | 12000<br>(7000,15000) | 11000<br>(6000,14000) | 12000<br>(7000,15000) | 10500<br>(6000,15000) | - | 0.707 |
| IL-6 (pg/ml) | 23.6<br>(17.5,43.8) | 21.7<br>(17.5,43.8) | 19.7<br>(16.4,28.7) | 19.7<br>(16.7,32.4) | - | **0.001** |
| Procalcitonin<br>(ng/ml) | 0.5 (0.3,0.65) | 0.54 (0.3,0.7) | 0.45<br>(0.23,0.67) | 0.5<br>(0.23,0.65) | - | 0.348 |
| D-dimers<br>(mcg/ml) | 15 (8,32) | 15 (7,33) | 15 (5,28) | 21.5 (9,34) | - | 0.556 |
| CRP (mg/dl) | 17 (11,54) | 23 (9,60) | 17 (9,54) | 24 (9,49) | - | 0.146 |

**Figure 1.**
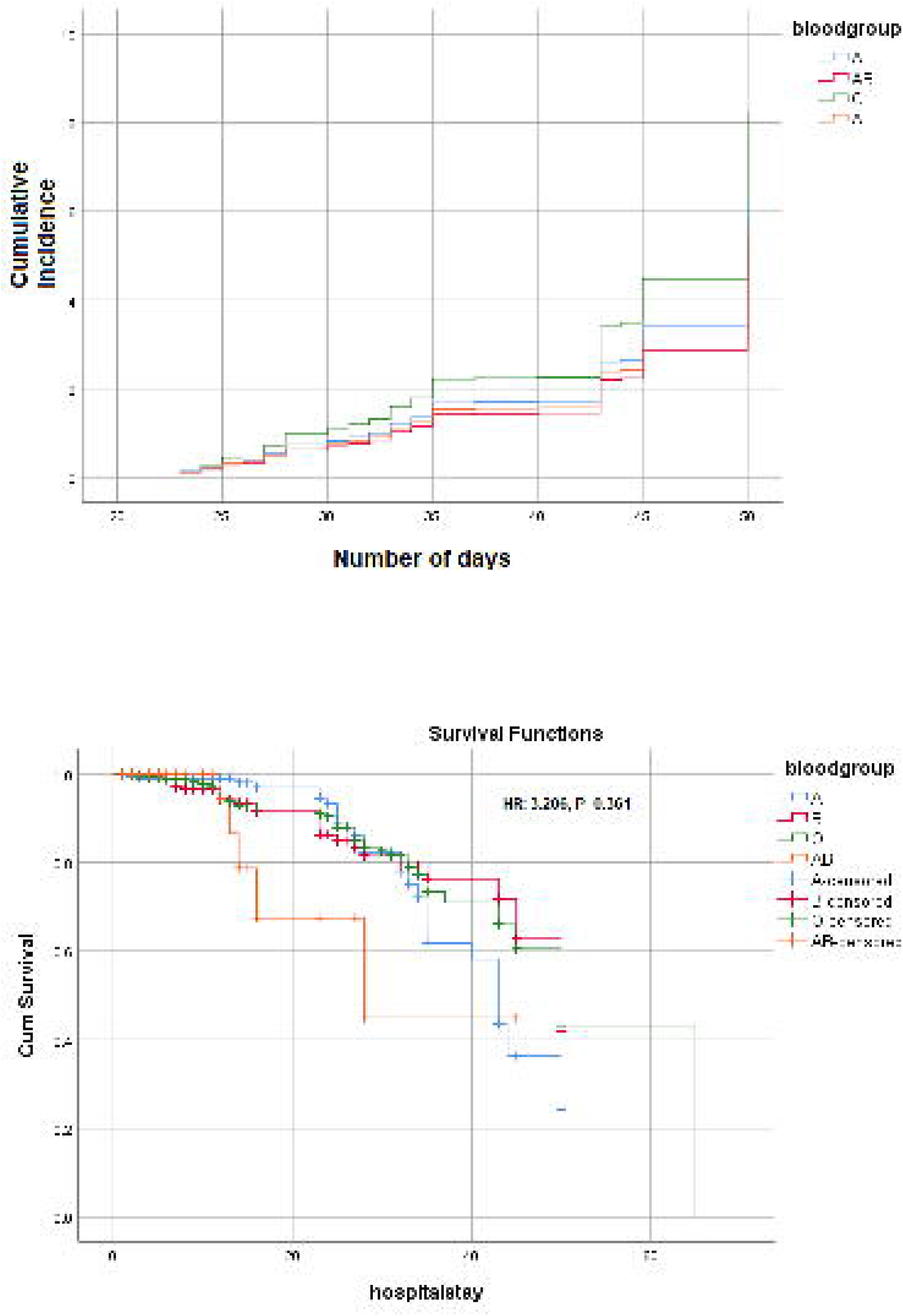
Cumulative incidence and survival function Kaplan Meier curve of ABO blood type in COVID-19 cohort.

**Figure 2.**
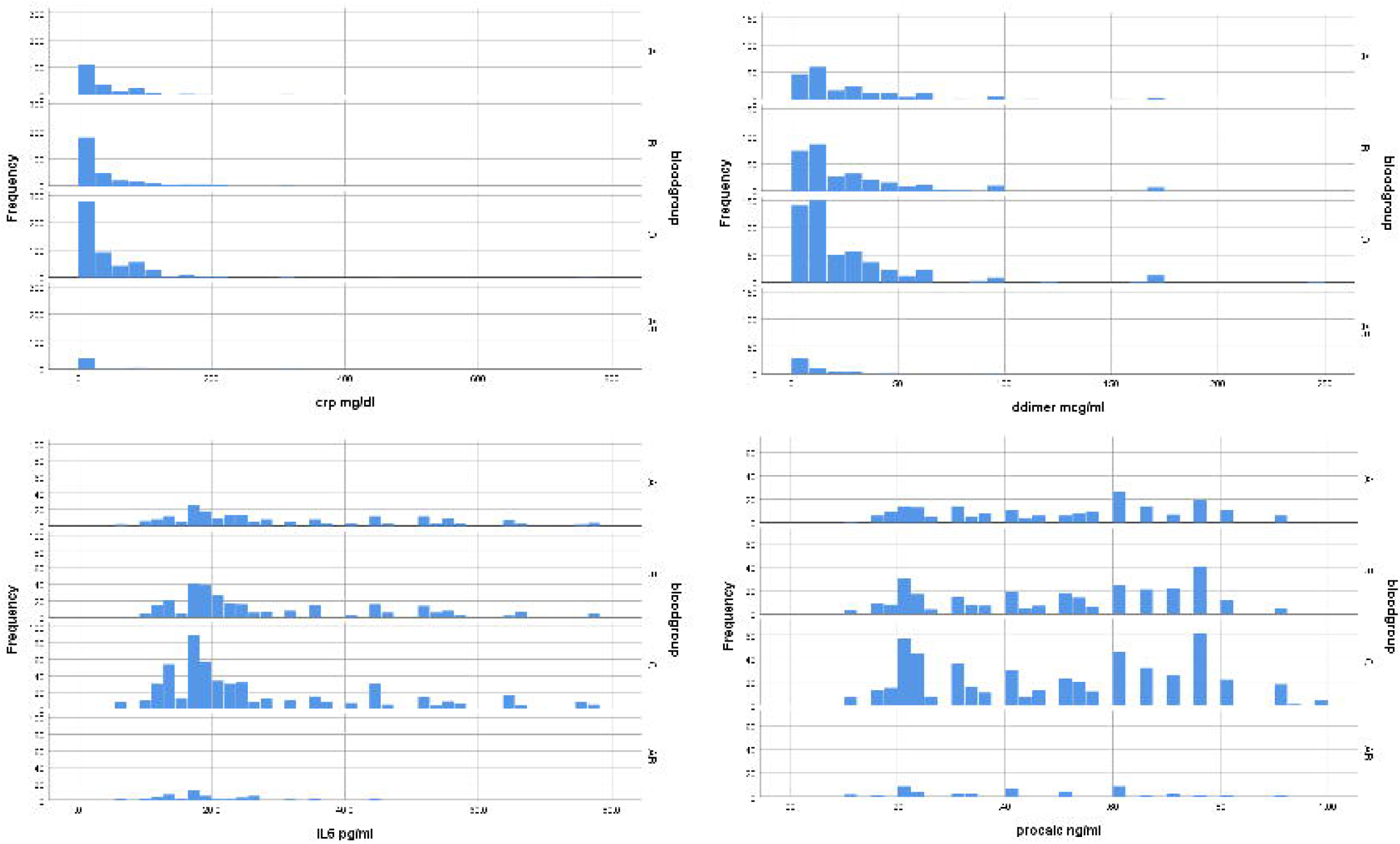
Frequency distribution of acute phase reactants in correlation with ABO blood groups.

Logistic regression demonstrates blood group O and AB as predictors for severe COVID-19 disease and mortality (O: OR: 0.438 (95% CI: 0.168-1.139) p=0.090; AB: OR: 0.415 (95% CI: 0.165-1.046) p=0.062) in this cohort (**Table 2**). The cause-specific hazards ratio (HR) for survival function was 3.206 (p=0.361) among all blood groups (**Figure 1**). More survival was seen with blood group A initially but with a hospital stay of more than approximately 30 days, the survival decreased exponentially in blood group A, while early mortality was observed with blood group AB (**Figure 1**).

**Table 2.**
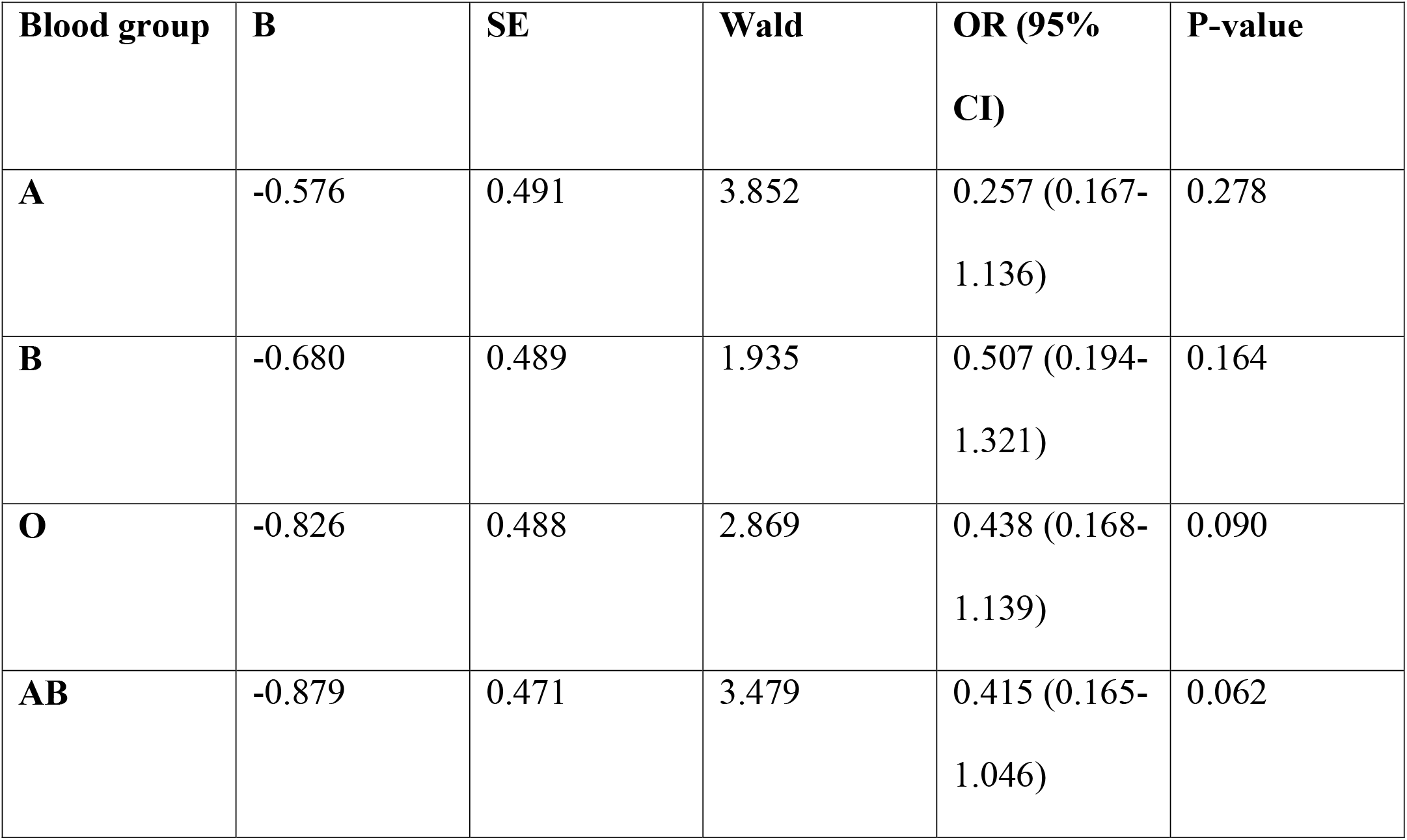
Logistic regression analysis on ABO blood groups showing predictors of severe COVID-19 disease. Odds ratio (OR), confidence interval (CI). P<0.05 as significant.

| Blood group | B | SE | Wald | OR (95% CI) | P-value |
| --- | --- | --- | --- | --- | --- |
| **A** | -0.576 | 0.491 | 3.852 | 0.257 (0.167-1.136) | 0.278 |
| **B** | -0.680 | 0.489 | 1.935 | 0.507 (0.194-1.321) | 0.164 |
| **O** | -0.826 | 0.488 | 2.869 | 0.438 (0.168-1.139) | 0.090 |
| **AB** | -0.879 | 0.471 | 3.479 | 0.415 (0.165-1.046) | 0.062 |

## Discussion

Of all the human blood group systems, the most widely used in clinical practice is the ABO blood group and includes four blood types, including A, AB, B, and O. It is located on chromosome 9 in a human DNA (9q34.2) and many studies have demonstrated a vital role of the ABO blood group in some infectious and non-infectious diseases. Histo-blood group antigens (HBGAs) are one of the main antigens expressed on human red blood cells, and differences in blood group antigens can alter host susceptibility to many infections. HBGAs are postulated to decrease the spread of infections through antibodies and ABO antibodies are a part of the innate immune system against many pathogens [10].

In this study, we demonstrated a relationship of COVID-19 severity with ABO blood groups. Blood groups A and O had the majority of severe cases in this cohort of COVID-19 patients. By contrast, we observed mild disease to be prevalent in the B and AB blood groups. Acute phase reactants were majorly deranged in blood group O as compared to other groups and hospital stay was associated more with A and AB blood groups. Overall mortality was high at 10.5% and blood group A was associated with the highest COVID-19-associated mortality. The majority of blood group reported in this cohort was O (48.8%).

There are several studies demonstrating a relationship between ABO blood groups and COVID-19 disease. A study from China tested the association of ABO blood group with COVID-19 infection on 105 COVID-19 cases and 103 controls. Blood group A was prevalent in their population (42.8%) and it was statistically associated with increased infection risk of COVID-19 (OR: 1.33, 95% CI: 1.02-1.73, p=0.04) in their female population [11]. Two recent studies from the subcontinent: one from Peshawar, Pakistan, and the other from Dhaka, Bangladesh have demonstrated the susceptibility of COVID-19 with the ABO blood groups [12,13]. The study from Pakistan had a sample size of 1935 patients, with blood group B as the prevalent blood type (35.9%) with an increased susceptibility for COVID-19 infection (OR: 1.195 (95% CI: 1.04-1.36), p=0.009) while blood groups A and O did not have statistically significant association of positive RT-PCR for SARS-COV-2. The other study from Bangladesh included 381 patients with a prevalence of blood group A in the COVID-19 cohort (32.9%, p<0.001), and no significant differences were observed in the duration of symptoms among other blood types. Our study exhibits contrasting results when compared to these investigations. Blood group O was prevalent in our study cohort (48.8%) and severe disease was associated with blood group A and O (41.1%, 35.1%, respectively). Hospital stay was more with blood type A and AB and higher mortality was associated with blood type A as well (13.9%).

In our previous study and a preprint, we demonstrated altered lipid profiles and thyroid function tests in association with COVID-19 disease and its severity along with acute phase reactants [14,15]. Acute phase reactants were severely deranged in both study cohorts and there was a positive correlation with the higher classification of severity of COVID-19. A similar conclusion was given in a systematic review of 34 articles, showing the derangement of laboratory parameters with increasing severity of COVID-19 [16]. In this investigation, blood group O was linked to severe derangement of acute-phase reactants even though blood type A included a majority of severe cases. In contrast, a study from Canada reported no difference in acute phase reactants between blood groups A, AB, O, or B [17]. This phenomenon is still unclear with heterogeneous results in different populations and needs further work to understand the underlying mechanisms.

There were several limitations to this study. First, it was a single-center study and the retrospective nature limits the control of confounding factors. Second, due to the limited sample size of COVID-19 in the early stages, the sample size included in this study was not very large. Third, as it represented the population of one province, so a regional selection bias needs to be considered. Third, other comorbidities might influence the severity of the disease.

## Conclusion

In conclusion, we demonstrate that male patients with blood group A and O are associated with an increased risk of severe COVID-19 disease after gender stratification while group A is associated with increased hospital stay and mortality. Alteration of the acute phase reactants is positively associated with the most prevalent blood type in this cohort, the blood type O.

## Data Availability

Available upon request

## Notes

### Competing Interest Statement

The authors have declared no competing interest.

### Funding Statement

None to declare

### Author Declarations

Ethical review board of Advanced diagnostics and liver center approved the study after review. Written/ informed consent was taken from all the participants/guardians before data extraction. (ID: ADC/17/20)

